# Impact of visual impairment on balance and visual processing functions in students with special educational needs

**DOI:** 10.1101/2020.09.28.20202879

**Authors:** KY Choi, HY Wong, HN Cheung, JK Tseng, CC Chen, CL Wu, H Eng, GC Woo, AMY Cheong

## Abstract

**Introduction:** Vision is a critical factor for children’s development. However, prevalence of visual impairment (VI) is high in students with special educational needs (SEN). Other than vision disability, this group of students is prone to having functional deficits. It is unclear whether visual problems relate to these compromised functional deficits. This study aimed to assess the impact of vision on visual processing functions and balance performance in SEN students through a community service in special schools.

**Methods:** A total of 104 (chronological age 14.3 ± 4.3 years, 43 females) SEN students in Taiwan were assessed and classified as having normal vision (NV) or vision impairment (VI). Visual acuity (distance and near) and contrast sensitivity (CS) were measured as the visual outcomes. Visual processing function assessment included facial expression recognition by Heidi expression test, in terms of card matching (FEC), and examiner’s facial expression matching (FEE), and visual orientation recognition (by mailbox game, VO). Dynamic balance was assessed with Timed Up and Go (TUG) test, while static standing balance was assessed using a force plate to measure the postural sway in double-legged feet-together and tandem stance with eyes open and closed conditions. Static balance was presented in terms of the change in the centre of pressure in maximal medial-lateral (ML) and antero-posterior (AP) sways, sway variability (V), and sway path length (L).

**Results:** Although visual acuity was significantly worse in VI than NV (p < 0.001), CS was similar in the two groups (p = 0.08). VO, FEC, and FEE also did not differ significantly between groups (p > 0.05). NV performed better in the TUG than VI (p = 0.03). There was a significant interaction between eye condition and the vision group (p < 0.05) for static balance. Pairwise comparisons showed that NV swayed significantly less in ML than VI under tandem stance-open eye condition (p = 0.04), but significantly more in closed eye condition (p = 0.03). Conversely, VI had less V and shorter L than NV under tandem stance-closed eye condition (p = 0.03).

**Conclusion:** This study is the first to our knowledge to examine the effect of vision on visual processing functions and balance performance in SEN students. Vision did not appear to be the major reason for impairment in visual processing. However, vision plays an important role in maintaining dynamic and static balance in SEN students.

## Introduction

Vision in children is crucial for their daily life, as it contributes greatly to the development of their functional abilities and essential skills needed for schooling and learning, such as reading comprehension and mathematical concepts. However, visual disabilities are common in children with special education needs (SEN).^1^ Perinatal adversity is one of the major causes of vision loss,^2, 3^ including preterm birth, improper neonatal environment, and neurological damage, which can affect visual acuity, contrast sensitivity, and ocular alignment. The level of visual impairment has been shown to be dependent on the severity of the SEN (e.g. grading of cerebral palsy),^4^ which was partially attributable to cerebral visual impairment.^5^ In addition to neurological and anatomical damage, correctable refractive error was also highly prevalent in children with SEN,^6, 7^ so prescription of spectacles or other optical aids would be beneficial to improve their vision. However, recent evidence has indicated inadequate eye care service for children with SEN,^8, 9^ leading to an inappropriate educational experience due to the misunderstanding of the individual’s visual status.

Balance function is also compromised in children with SEN, in terms of static and dynamic balance. Children with more severe cerebral palsy (higher gross motor function classification system level) recorded a greater magnitude of postural sway during a static balance measure than children with normal development,^10^ as well as dynamic balance function measured with the Timed Up and Go (TUG) test.^11^ Children with Down syndrome tended to spend more time to execute functional balance tasks, such as performing a standing reach^12^ and the TUG test.^13^ The postural control system was also found to be underdeveloped in children with autism spectrum disorder (ASD)^14^ and deafness.^15^ For children with other intellectual disabilities, it was controversial whether their balance function was inferior to their peers with normal development.^16, 17^ However, visual impairment was associated with reduced balance function.^17, 18^ It was also reported that postural sway for male participants with visual impairment, regardless whether their eyes were open or closed, was similar to that of sighted participants with their eyes closed.^19^ This implied that vision had less impact on postural control despite compromised balance function in visually impaired patients. Balance function in children with disruption of binocular vision, due to strabismus or amblyopia, was significantly reduced.^20^ However, postural control was significantly improved after corrective surgery in children with strabismus,^21^ suggesting that there was a possibility to improve balance function by correcting vision or improved visual function. Children with visual impairment had poorer postural sway in both double-leg and single-leg standing compared with their sighted peers.^22^ However, reduced static balance function was only found in eye-open, but not eye-closed condition. Both dynamic functional balance and coordination were also reported to be weaker in children who were visually impaired.^23, 24^ Despite the important role of vision on balance function, no studies have considered the impact of visual impairment in children who are more prone to have compromised balance function (e.g. children with SEN).

Visual processing is the ability of the brain to acquire, compute, and interpret visual information. In view of the importance of vision in childhood development, thorough assessment of visual function and related visual processing functions can provide information on the limitations of the SEN children.^25^ Visuo-spatial and visuo-perceptual impairment, in terms of facial recognition and line orientation judgement, are common in children with bilateral cerebral palsy^26^ and ASD.^27, 28^ Extremely pre-term birth or very low birth weight were associated with visual processing disorders and poorer academic performance in adolescents, after controlling for other perinatal risk factors.^29^ However, based on the various capabilities of the SEN children, it has been suggested that evaluation for visual processing function could be better delivered in play situations,^30^ especially for children who functioned normally in other visual tasks, such as visual acuity.^31^ Despite the importance of vision on learning different perceptual skills, it is unclear whether visual impairment further impairs visual processing performance in children with SEN.

Previous studies have suggested poorer balance function, weaker visual processing function, and higher prevalence of vision impairment in students with SEN. However, it is unclear whether compromised vision further impairs these SEN students’ balance and visual processing performance. To our knowledge, this is the first study aiming to investigate the impact of vision on visual processing function and balance performance (dynamic and static) in students attending special-care schools in Taiwan.

## Methods

### Study population

This study analysed cross-sectional data collected as part of a community service project to provide eye care services for students with special needs attending ten special schools in Taiwan. All tests were conducted by optometrists and trained university students. Referring to the study designed by Klavina et al.,^18^ to detect a difference in postural balance between visually impaired and intellectually disabled students, 20 students in each group were needed to detect an effect size of 1.20 (GPower, two-tail test, α = 0.05, Power = 95%). Given the wide variety of SEN students recruited, a more conservative effect size of 1.0 was adopted, with a sample size of 29 in each group. A total of 157 students agreed to participate in the study, of whom 127 had measurable visual acuity and at least one functional measurement. Students with solely visual impairment, but otherwise normal development (i.e. absence of other non-visual disabilities, n = 23) were excluded. The remaining 104 students (chronological age 14.3 ± 4.3 years, range 4 – 19 years, 43 females) were included in the analysis to assess the impact of vision on students with different disabilities, including cerebral palsy, ASD, Down syndrome, and other intellectual disabilities. Written consent and verbal assent (if feasible) were obtained from the guardians and the students, respectively. All study procedures followed the tenets of the Declaration of Helsinki and were approved by The Hong Kong Polytechnic University Human Subjects Ethics Subcommittee.

### Data collection

Demographics and information on the disabilities were obtained by a structured questionnaire completed by either the guardians or school teachers. The questionnaire included birth history, types of disabilities, self-report vision status, and visual problems. The subtypes of SEN were classified as cerebral palsy (CP), ASD, Down syndrome (DS), isolated intellectual disability (ID), and Others which included deaf-mute, Rett syndrome, inborn errors of metabolism, etc.

Distance visual acuity was measured monocularly by Lea symbols at 3 m (or 1.5 m if vision was poor) using matching toys (The Good-Lite Company, USA). If the students were intellectually incapable of performing the test, tests with lower cognitive requirement were used, sequentially Cardiff acuity test at 1 m by pointing at the direction, Cardiff acuity test at 1 m by preferential looking, and Lea gratings at 57 cm by preferential looking. Habitual visual acuity of the better-seeing eye was converted into LogMAR acuity, allowing subjects to be classified into two groups: normal vision (NV, LogMAR < 0.50) and visual impairment (VI, LogMAR ≥ 0.50).

Habitual visual acuity was used for grouping because the functional performances were assessed with visual correction aids. Near visual acuity was measured binocularly by LEA near vision card (The Good-Lite Company, USA). Binocular contrast sensitivity (CS) was measured using the letter version of Mars (The Mars Perceptrix Corp., US), or Lea low-contrast symbols flip-chart test / Hiding Heidi test (The Good-Lite Company, USA) at 40 cm if students were intellectually incapable, then LogCS was recorded. External and internal ocular health was assessed using slit lamp biomicroscopy and direct ophthalmoscope (or binocular indirect ophthalmoscope with pupil dilation upon parent’s consent), respectively.

Visual processing function was evaluated by two paediatric tests (The Good-Lite Company, USA). Visual orientation recognition was measured using the Lea mailbox game, in which the students were asked to drop a card through a slit opening in different orientations, and the average time for five trials was recorded (VO). Facial expression recognition was measured using the Heidi expression test. The test was divided into two parts: (1) expression recognition with the Heidi cards matching (FEC): students were given a set of expression cards, then they were asked to match the card with that the examiner displayed, (2) expression recognition with the examiner’s facial expression (FEE): students were given a set of expression cards, then they were asked to match the facial expression that the examiner expressed on his/her face. Both parts were timed, and the average time needed for the five trials was recorded.

Dynamic balance function was measured by the Timed Up and Go (TUG) test.^32^ The students were asked to rise from a chair, walk three meters on a straight line, turn around, return to the chair, and sit down. The chair height depended on the height of the students using the common chairs provided by the schools, as the chronological age of the sample covered a wide range. The test was repeated three times, with the time needed for each trial being averaged and recorded.

Static balance function, in terms of postural sway, was measured using a force plate (BP400600, AMTI, US) with double-leg feet-together standing and tandem stance. Double-legged feet-together standing was chosen to mimic the natural standing position in daily life, while tandem stance condition was chosen over one-leg standing to mimic the postural stability during adverse conditions because of the limited capability of the SEN students in the current study. Postural sway was measured at each condition while standing steadily for 20 s under each of the four conditions: (1) feet together + eyes open (FO); (2) tandem stance + eyes open (TO); (3) feet together + eyes closed (FC), and (4) tandem stance + eye closed (TC). Students were asked to fixate at a distant target at 3 m under eye open condition (Figure 1). The force plate measured the subjects’ position at the centre of pressure (COP), at which several variables were generated and included in the analysis: maximum medial-lateral and antero-posterior sway (ML and AP – i.e. maximal amplitude of COP in the ML and AP dimensions), sway variability (V – i.e. variability of COP around its mean value computed by the root mean square of the COP displacement in ML and AP), and sway path length (L – i.e. total distance of COP travelled).

**Figure 1.**
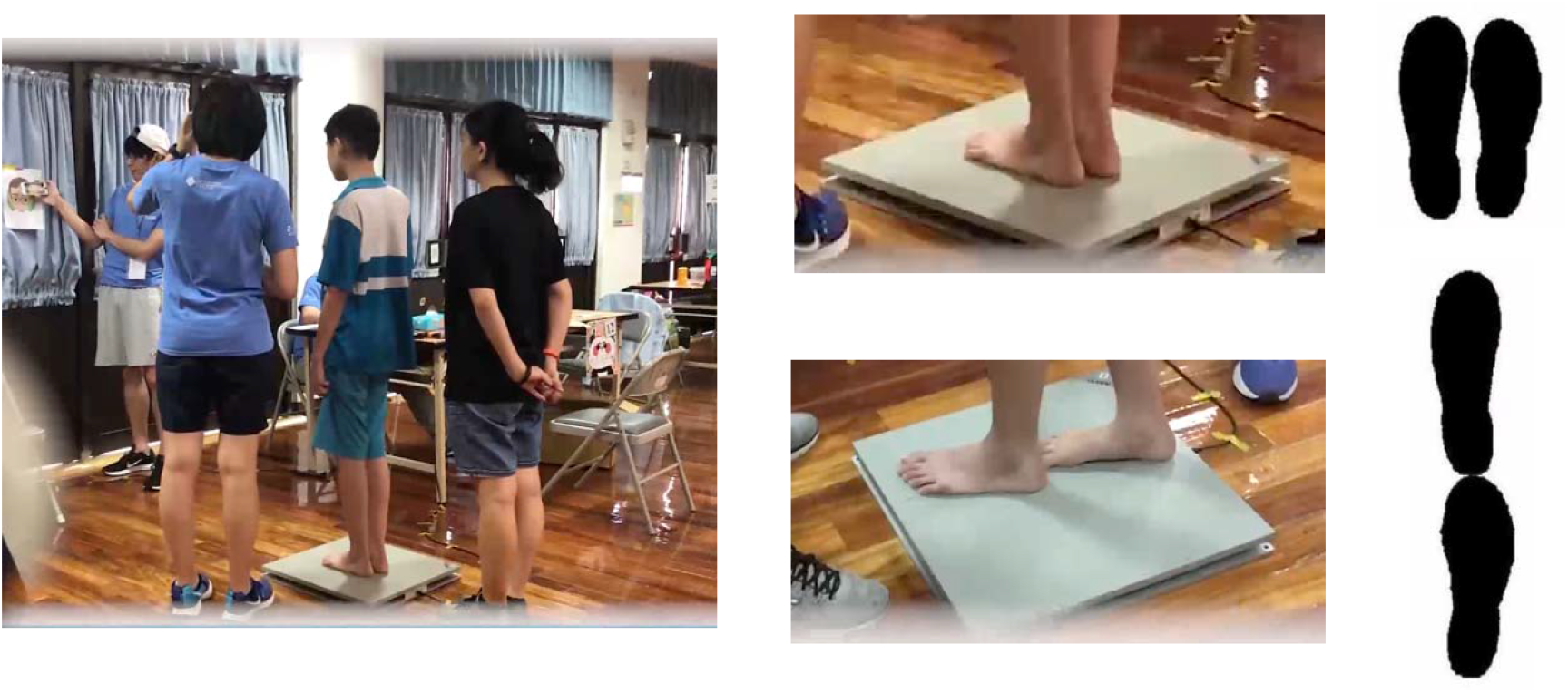
Measurement of postural sway while standing on a force platform for different conditions. Students were asked to fixate at a distant target at 3 m under open eye condition (left panel). Upper condition of the right panel was double-leg standing with feet together, while the lower condition was tandem stance.

### Statistical analysis

As the distribution of some of the outcome variables was significantly different from normal, non-parametric tests were used in the analysis. Chi-square (*χ*^2^) test was used to evaluate the distribution of visual impairment among the subtypes of SEN. Kruskal-Wallis test and Mann-Whitney U test were used to compare visual functions, visual processing functions, and dynamic balance function between the two vision groups. Comparison among the subtypes of SEN is included in Supplementary Table 1. For static balance, the results were transformed to achieve normality using percentile ranking followed by inverse-normal transformation into normally distributed Z-scores.^33^ Two-way repeated measures ANOVA was used to analyse the main effects and the interaction effect between the grouping (NV and VI), eye conditions, and feet conditions on the static balance. Greenhouse-Geisser test was used whenever sphericity could not be assumed, and Bonferroni adjustment was used in the post-hoc comparisons. Inability to perform the test was regarded as missing data, which was treated with pairwise deletion. Static balance was not compared among subtypes of SEN because of the limited number of students who successfully completed different conditions in each group. Significance level was set as p < 0.05. All statistical procedures were performed using SPSS v22 (IBM Inc, US).

## Results

Of the 104 students, 62 (59.6%) were classified as having visual impairment (VI, i.e. visual acuity of the better eye was ≥ 0.50 LogMAR), in which the major causes of the reduced vision were optic nerve related (18.8%, including optic atrophy, optic nerve hypoplasia, and glaucoma), retinal impairment (17.6%, including retinopathy of prematurity, retinal dystrophy, and macular anomalies), uncorrected/under-corrected refractive error (14.1%), ocular media opacity (12.9%), and oculomotor anomaly (11.8%). However, a large proportion of VI (24.8%) was probably not due to ocular problems, but other neurological causes (e.g. cerebral visual impairment, which refers to the vision loss caused by retro-geniculate damage in the absence of ocular abnormalities). Only 23 (22.1%) of the 104 students had optical correction (11 NV, 12 VI) before participating in this study. After subjective refraction or retinoscopy, 32 students (30.8%, 12 NV and 20 VI) were found to have uncorrected (24) / under-corrected (8) refractive errors and benefited from prescription of updated optical aids to improve their vision thereafter, with a mean improvement of visual acuity of LogMAR 0.33 ± 0.17. Twelve out of 20 VI students, who were prescribed with an updated optical aid, had best-corrected visual acuity better than LogMAR 0.50, reducing the prevalence of VI by 11.5%. Notably, among the 62 VI students, only 25 guardians/ teachers (40.3%) reported that they were aware of visual problems in their children/ students in the questionnaire, revealing an insufficient awareness of visual problems encountered in the SEN population. Given the different capabilities of the students, it was expected that some students could not complete all the tasks. The distribution of missing data in visual processing and balance functions were independent of the grouping (p > 0.05), indicating a similar capability in two groups. The demographics, visual functions, visual processing functions, and balance performances in terms of vision groups are listed in Table 1, whereas those in terms of the subtypes of SEN are listed in Supplementary Table 1.

**Table 1.** Comparison of demographic, visual and functional performance between vision groups [median (IQR) or percentage]

|  | Normal vision<br>(NV, n = 42) | Visual<br>impairment<br>(VI, n = 62) | p-value |
| --- | --- | --- | --- |
| Demographic: |  |  |  |
| Chronological age (in years) | 16.0 (5.3) | 15.0 (5.0) | 0.32 |
| Sex |  |  | 0.88 |
| Female | 17 (40.5%) | 26 (41.9%) |  |
| Gestation age |  |  | 0.78 |
| Full-term ($\geq 37$ weeks) | 28 (66.7%) | 42 (67.7%) | |
| Pre-term ($< 37$ weeks) | 6 (14.3%) | 11 (17.8%) | |
| Unknown | 8 (19.0%) | 9 (14.5%) |  |
| Self-reported visual disabilities | 6 (14.3%) | 25 (40.3%) | **< 0.01** |
| Vision measures: |  |  |  |
| Distance acuity of the better eye (LogMAR) | 0.30 (0.26) | 0.87 (0.55) | **< 0.001** |
| Near acuity (LogMAR) | 0.21 (0.35) | 0.65 (0.89) | **< 0.001** |
| Contrast sensitivity (LogCS) | 1.64 (0.30) | 1.60 (0.86) | 0.08 |
| Functional measures: |  |  |  |
| Visual orientation (VO) – time (s) | 1.68 (1.37) | 1.78 (1.42) | 0.71 |
| Facial expression (Cards, FEC) – time (s) | 11.00 (13.25) | 15.34 (19.90) | 0.14 |
| Facial expression (Examiner, FEE) – time (s) | 15.00 (25.33) | 18.00 (36.30) | 0.52 |
| Time-up-go (TUG) – time (s) | 10.60 (4.52) | 13.61 (9.05) | **0.03** |
| Static balance (medial-lateral sway, ML) (cm)† |  |  |  |
| (i) Feet-together with eyes open (FO) | 1.62 (0.93) | 1.87 (1.05) |  |
| (ii) Tandem stance with eyes open (TO) | 1.92 (0.68) | 2.45 (0.91) |  |
| (iii) Feet-together with eyes closed (FC) | 1.47 (1.23) | 1.81 (1.50) |  |
| (iv) Tandem stance with eye closed (TC) | 2.69 (0.97) | 2.23 (1.39) |  |
| † Only 20 (47.6%) and 32 (51.6%) students from the NV and VI groups, respectively, could complete the different conditions in the static balance. |  |  |  |

In terms of vision, the chronological age distribution was similar in NV and VI groups (p = 0.44) and the LogCS did not differ significantly between two groups (Mann-Whitney U = 886.00, p = 0.08). Chronological age was not significantly associated with performance in visual processing functions (Spearman’s test, p > 0.05). VI and NV groups also had similar performances for the visual processing functions, in terms of VO, FEC, and FEE (p > 0.05). With respect to dynamic balance, NV students performed significantly better than VI students (Mann-Whitney U = 1040.00, p = 0.03) by a median difference of 3.61 s to complete the TUG task.

The results for different balance variables among vision groups are shown in Figure 2. In general, SEN students’ static balance function varied substantially when measured in different conditions. Results from the two-way repeated measures ANOVA showed that the within-subject main effects for eye condition (i.e. eye open vs. eye closed) and feet condition (i.e. feet together vs. tandem stance) were insignificant (F_1,50_ < 1.15, all p > 0.05) in all the outcomes, as was between-subject effect for vision group (i.e. NV vs. VI) (F_1,50_ < 0.92, all p > 0.05). However, the interaction between the eye condition and vision group was significant in V (F_1,50_ = 4.24, p = 0.05) and L (F1,50 = 4.66, p = 0.04), revealing that the absence of visual input (i.e. eye closed condition) had stronger impact on balance function in students with NV than those with VI. Further interaction among all three factors (eye condition, feet condition, and vision group) was significant in the ML (F_1,50_ = 7.44, p = 0.01), but not in other outcomes (p > 0.05). Detailed statistical results are listed in Table 2.

**Table 2.** Results of two-way mixed repeated measures ANOVA for static balance measures.

|  | F<sub>1,50</sub> | p-value |  | F<sub>1,50</sub> | p-value |
| --- | --- | --- | --- | --- | --- |
| <u>Medial-lateral sway (ML)</u> |  |  | <u>Antero-posterior sway (AP)</u> |  |  |
| Group | 0.92 | 0.34 | Group | 0.18 | 0.67 |
| Eye condition | < 0.001 | > 0.99 | Eye condition | 0.08 | 0.77 |
| Feet condition | 0.001 | 0.97 | Feet condition | 1.15 | 0.29 |
| Eye * Group | 2.92 | 0.09 | Eye * Group | 0.34 | 0.57 |
| Feet * Group | 0.33 | 0.57 | Feet * Group | 3.37 | 0.07 |
| **Eye \* Feet \* Group** | **7.44\*\*** | **0.01** | Eye * Feet * Group | 1.85 | 0.18 |
| <u>Sway variability (V)</u> |  |  | <u>Sway path length (L)</u> |  |  |
| Group | 0.61 | 0.44 | Group | 0.50 | 0.48 |
| Eye condition | 0.21 | 0.65 | Eye condition | 0.09 | 0.77 |
| Feet condition | 0.06 | 0.81 | Feet condition | 0.02 | 0.89 |
| **Eye \* Group** | **4.24** | **0.05\*** | **Eye \* Group** | **4.66** | **0.04\*** |
| Feet * Group | 1.25 | 0.27 | Feet * Group | 1.46 | 0.23 |
| Eye * Feet * Group | 2.22 | 0.14 | Eye * Feet * Group | 2.14 | 0.15 |

\* indicates $p < 0.05$ , \*\* indicates $p < 0.01$

**Figure 2.**
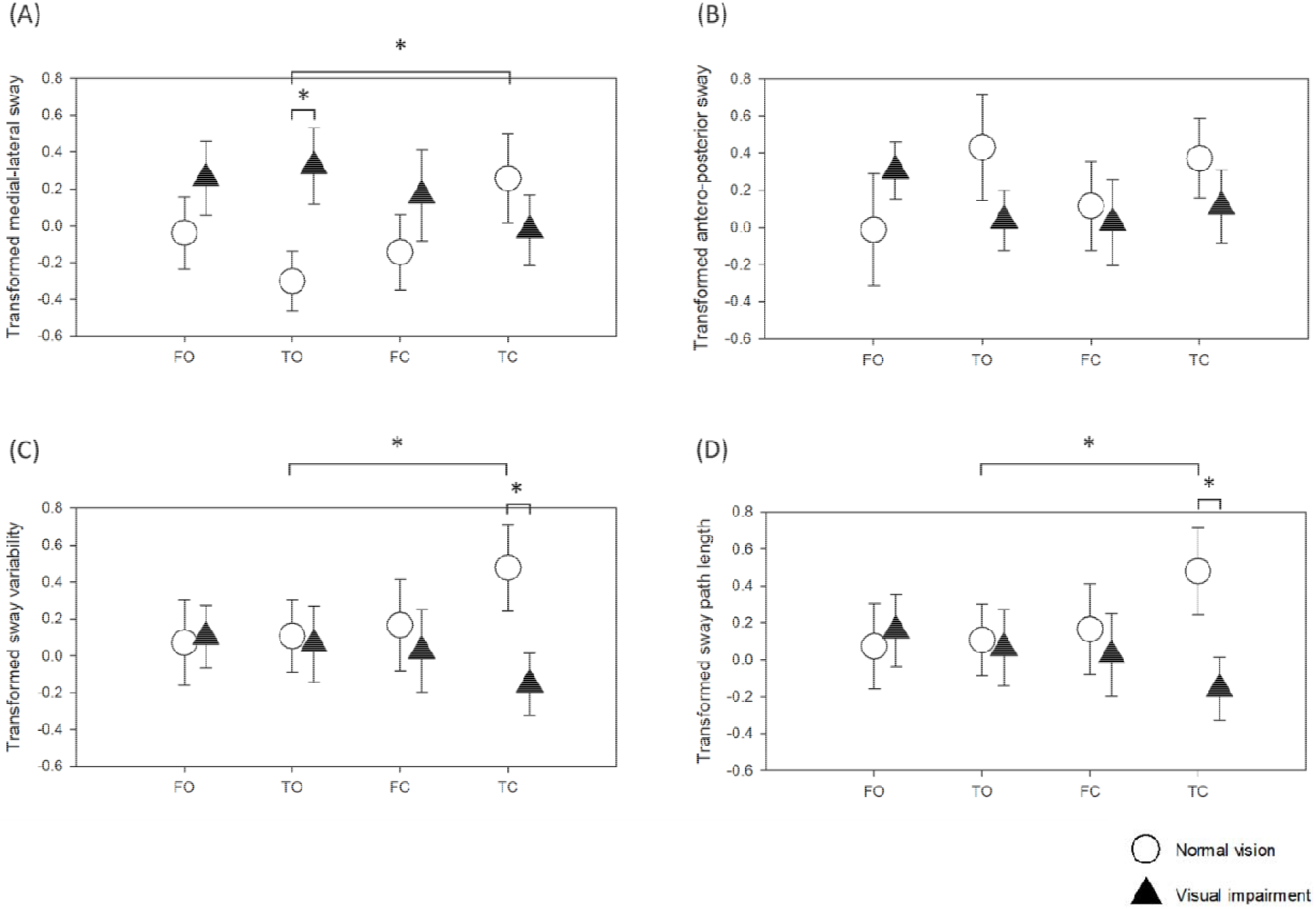
Static balance sway parameters in different eye and feet conditions of normal (NV) and vision impairment (VI) groups. (A) Medial-lateral sway (ML); (B) Antero-posterior sway (AP); (C) Sway variability (V); (D) Sway path length (L). FO: Feet together standing with eyes open; TO: Tandem stance with eyes open; FC: Feet together standing with eyes closed; TC: Tandem stance with eye closed. The graph is plotted with transformed data. The error bars represent the standard error of the mean.

In post-hoc pairwise comparisons, NV students swayed significantly less in ML than VI students under TO condition (p = 0.04), but ML sway became significantly greater under closed eye condition (p = 0.02). Similar findings were observed for V and L with NV students having significantly less sway under closed eye than open eye condition (p = 0.03). In contrast, VI students swayed significantly more than NV students in V and L under eye closed and tandem stance condition (p = 0.03). The findings demonstrated that when visual input was provided (i.e. open eye condition), students with normal vision had better static balance performance. However, when visual input was deprived (i.e. closed eye condition), students with vision impairment swayed significantly less.

Given the wide spectrum of disabilities in SEN students among different subtypes of SEN, further analyses were conducted. Chronological age differed significantly among the various subtypes of SEN (H = 16.21, p < 0.01). Students with SEN categorised as “Others” were significantly younger than those with DS (p = 0.02) and ID (p = 0.04), respectively by a median of 5.5 and 5.0 years. The habitual visual acuity and contrast sensitivity were similar among the subtypes of SEN (p > 0.05), but near visual acuity was better in Others (p < 0.01). However, students with different subtypes of SEN had significantly different performances in all visual processing function tests. For VO (H = 10.04, p = 0.04), students with Others performed significantly faster than those with ID (p = 0.03). These students also performed significantly faster than those with ID (p = 0.01), ASD (p = 0.01), and CP (p = 0.001) for FEC (H = 19.17, p = 0.001), and faster than those with ID (p = 0.01) and CP (p = 0.001) for FEE (H = 19.27, p = 0.001). Dynamic balance function also differed significantly among subtypes of SEN (H = 17.67, p = 0.001). Students with Others completed the TUG test faster than those with CP, DS, and ID (all p ≤ 0.01). Static balance function was not compared between SEN subtypes because of the limited number of students in each group. For example, only 2 of 13 ASD students could complete the static balance measurement due to difficulties in understanding or following the instructions.

## Discussion

This study is the first to our knowledge to examine the effect of vision on two essential daily tasks – visual processing and balance functions in children with SEN attending special education schools. In line with previous findings, prevalence of reduced visual acuity was high in this study. Our findings showed that VI (under current definition of LogMAR ≥ 0.5) group did not perform significantly worse in functional tests, such as orientation recognition and facial recognition – two important visual processing tasks, which could be due to the relatively low visual demand of the two functional tests. In contrast, vision was significantly associated with the balance function, both dynamic and static. SEN students with visual impairment had poorer performance in the TUG test and sway parameters under open eye condition. However, when the visual input was deprived (i.e. eye closed condition), students with normal vision performed significantly worse than those with visual impairment.

Previous studies have reported an increased risk, as high as 75%, of visual impairment in disabled children,^34^ whereas the current study found 59.6% of students attending the special schools had reduced vision, although this figure did not include those SEN students with solely visual impairment (n = 23). In the US, cerebral visual impairment was the leading cause of visual impairment and blindness in children^35, 36^ and constituted 19% of the visually challenged children. Our study, echoes the previous studies in finding that the causes of reduced vision for 24.8% of the SEN students were likely to be neurological, while the remainder were due to different types of ocular anomalies (e.g. optic atrophy, glaucoma). Although the types of disabilities were self-reported by guardians or teachers rather than full medical records (because of compliance with patients’ privacy),^37^ it was speculated that cerebral visual impairment was likely to be the major cause of vision loss in students attending the special schools.

Under the definition of LogMAR ≥ 0.5, VI did not appear to be a major obstacle for visual processing functions in the current study, which could be due to the inability of students to complete the required tasks and were excluded from the analysis (VO: n = 8; FEC: n = 24; FEE: n = 36). As in a previous study, visuo-spatial and visuo-perceptual abilities were found to be impaired in 90% and 60% of subjects with cerebral palsy, respectively,^26^ in terms of orientation judgement and facial recognition, which was independent of their visual acuity. Facial details processing deficits are also common in ASD,^27, 28^ even if these individuals have normal vision, leading to problems such as unsustainable eye contact and switching focus for social function. Inhibition of visual input caused by visual impairment might have limited impact on such social information. In addition, both sizes of the cards in the Lea mailbox game and Heidi expression test were large (10.2 x 10.2 cm), requiring low visual demand to complete the tasks. In the current study, VO, FEC, and FEE were similar in both NV and VI students. Similarly, chronological age did not show a significant association with the performance in visual processing functions (Spearman’s test, p > 0.05, Figure 3). In line with previous studies, our results indicated that non-visual disabilities, e.g. intellectual disability, might account for the impaired visual processing functions rather than vision itself. To facilitate the learning of the SEN students with VI, training tools of sufficient size should be employed to accommodate their visual needs, as our results indicate VI students had similar visual processing functions. Further studies on functional assessments, such as joint attention and imitation, are warranted. In addition, the current study focused on orientation judgement and facial expression recognition. However, other types of visual processing functions, e.g. visual discrimination, visual memory, spatial relations, visual-motor, and visual-auditory integration, are also critical for development in SEN students. Further exploration on the relationship between subtypes of SEN, visual functions, optical corrections, and visual processing functions would facilitate the design of training/learning modules and benefit the development of the SEN students.

**Figure 3.**
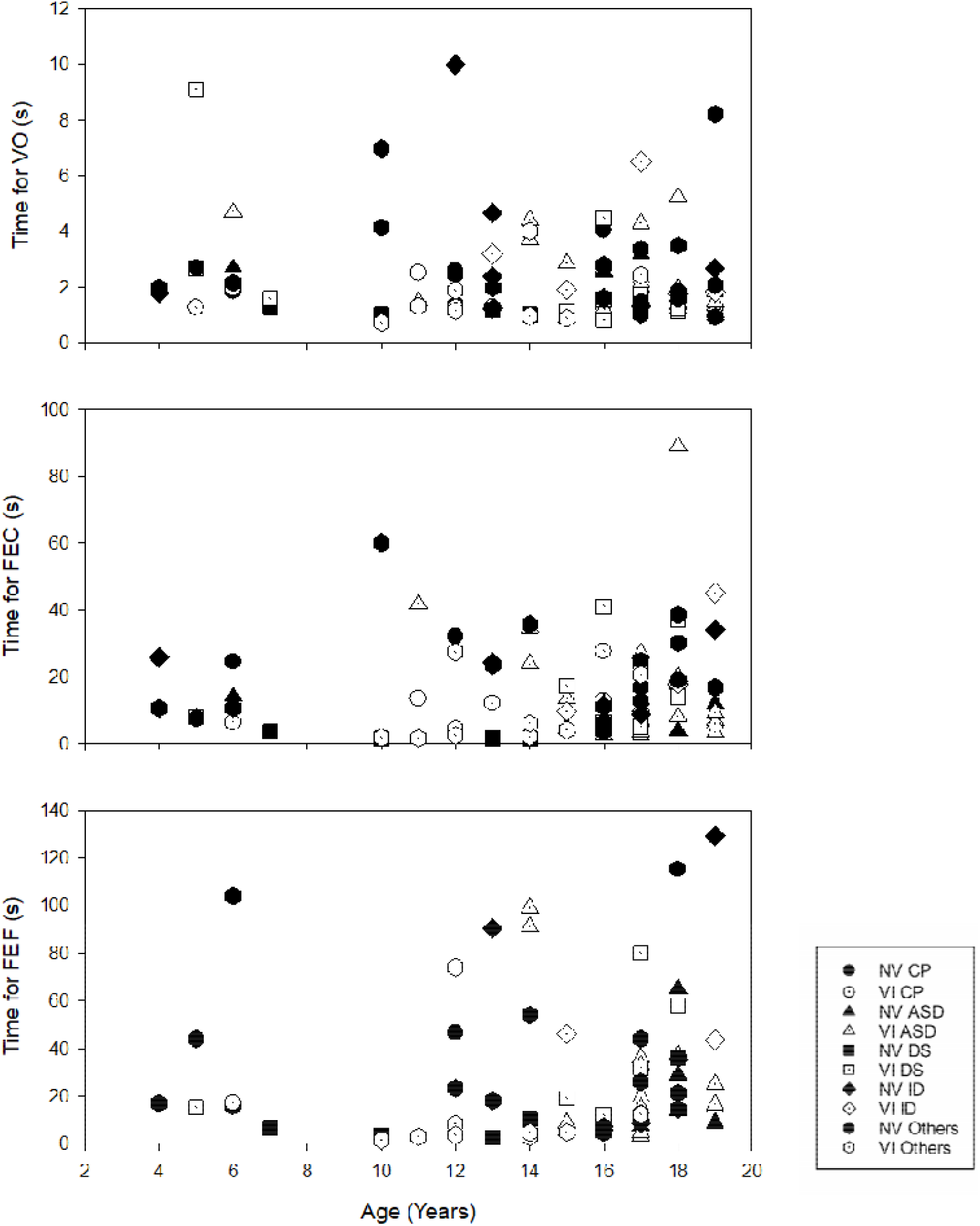
Relationship between visual processing functions and chronological age. Filled symbols indicate students with normal vision (NV), while dotted symbols indicate students with visual impairment (VI). Circle: Cerebral palsy (CP); Triangle: Autism spectrum disorder (ASD); Square: Down syndrome (DS); Diamond: Isolated intellectual disability (ID); Hexagon: Other disabilities.

The balance functions of SEN students are reduced, regardless of the type of disability. In children with normal development, the dynamic balance function measured by TUG ranged from approximately 4 to 7 s.^13, 38^ The time needed to complete the TUG task increased from a median of 7.5 s in gross motor function classification system level I, to 17.8 s in level II, and finally 50.7 s in level III for children with CP,^11^ while it was approximately 9 s in adolescents with DS.^13^ Our sample recorded a median of 10.60 s and 13.61 s in SEN students with NV and VI, respectively. The correlation analysis showed a significant, but weak association between habitual visual acuity and TUG (Spearman’s ρ = 0.23, p = 0.04). This indicated that visual and non-visual disabilities compromise SEN students’ dynamic balance function in different ways and might have a composite effect, as dynamic balance function was further reduced in students with both visual and non-visual disabilities. Several studies have compared static balance function in sighted and visually impaired participants. In summary, they concluded that visually impaired individuals had greater postural sway than sighted individuals under open eye condition, especially in single-leg-standing positions: adults,^19, 39^ adolescent,^17^ and children.^18, 22^ However, no significant difference was observed between the sighted and visually impaired individuals under closed eye condition. In the current study, a similar outcome was found in the SEN students. Those students with normal vision had better static balance performance in open eye condition than those with visual impairment under tandem stance, while no significant difference was observed in feet together standing. When visual input was deprived (i.e. eye closed condition), their static balance function became significantly poorer. However, students with both visual and non-visual disabilities (i.e. VI group) swayed similarly under eye open and eye closed conditions. It is speculated that students with visual impairment may rely more on their somatosensory and/or vestibular inputs rather than vision to maintain their postural control, while NV students rely more on the visual input. Coincidently, it was observed that similar balance performance was found in older adults with VI.^40^ They swayed significantly more than NV subjects in static standing under eye open, but displayed no difference with sighted aged-match subjects when eyes were closed. The reliance on somatosensory input was shown when subjects with VI stood under a sway-reference support surface with eye open swayed significantly more than the sighted subjects.^40^ Further studies examining the contribution of multi-sensory inputs to postural control, and functional activities, such as obstacle avoidance and crossing, are needed. Such knowledge could facilitate the design of balance training protocols for students with SEN.

Taiwan initiated a national-wide registry policy for children with disabilities in the 1980’s to better understand and provide support for those with SEN. The combined prevalence of SEN increased from 1.0% to 1.5% over three decades,^41^ affecting more males, especially in rural areas.^42^ Institutionalised care, e.g. special schools, has been the mainstream for the special care service,^43^ providing educational, occupational, and vocational training for registrants. Although resources had been pooled for rehabilitation services, the prevalence of the beneficiaries was still low,^44^ in which the service recipients only accounted for 24.5% of an 957-subject SEN sample within a 7-month period. As well as the students with SEN themselves, the primary family carer may also experience challenges, causing the carers’ health status and quality of life to be significantly lower.^45^ In the current study, despite the students attending special care schools, awareness of visual impairment of the guardians or teachers was still low, in that only 40.3% reported their children or students to have visual impairment. In addition, nearly one third of the SEN students (n = 32) were found to have uncorrected / under-corrected refractive error. Their visual acuity was significantly improved by an average of LogMAR 0.33 by merely updating their optical aids. This phenomenon indicates the necessity of regular eye examinations for the SEN population, as suggested in previous studies.^8, 9, 46^

There were several limitations in the current study. Firstly, the self-reporting disability status was less reliable than reviewing a full medical history. However, due to the compliance with patient privacy, this information could not be retrieved from the guardians or teachers. Secondly, the severity of non-visual disabilities was not assessed in the current study, which is speculated to affect the capability of SEN student to perform different tasks. Most of our tests required students to have a certain degree of cognitive ability to participate. It is not surprising that almost half of the participating students could not complete the various conditions in balance measurement, in particular the tandem stance in eye closed condition. Finally, this study lacked a longitudinal follow-up to observe the effect of improved vision after updated optical corrections on visual processing functions (both those measured in the current study and others, including reading speed, visual memory, and spatial relations) and balance functions in SEN students.

To conclude, visual impairment in SEN students is common. However, despite the high prevalence of visual impairment in this population, our findings suggest that some SEN students’ visual function could be improved by prescribing appropriate and updated refractive correction. Although slightly reduced vision is not a major limiting factor explaining the deficits of visual processing function in these SEN students, vision remains an important integral for children’s all-round development and parents and teachers could use tools of sufficient size for training and learning purposes to overcome their visual impairment. Adequate visual input was also found to be critical for SEN students to maintain their dynamic and static balance functions. Hence, regular checking and preserving of the vision of SEN students is of high importance.

## Data Availability

The data can be provided upon request.

## Acknowledgment

This study was supported by The Hong Kong Polytechnic University Community Service Fund (83CT) and Chinese Mainland Affairs Office Fund. We thank the financial support from Lions Clubs International MD300 Taiwan Eye Care Network Committee, Taiwan. We thank Dr Pei-Chang Wu and other ophthalmologists from Chang Gung Hospital for providing part of the ocular health examination during the trip. In addition, we appreciated the assistance provided by all volunteering students from The Hong Kong Polytechnic University and Asia University, volunteers from Taiwan Lions Club and teachers from the participating schools. We also thank Prof. William Tsang and Ms. Eva Chan from The Open University of Hong Kong to provide professional advice in preparation of this manuscript. We acknowledge Dr. Maureen Boost for proofreading this manuscript.

