## Supplementary Table 1 for "Impact of visual impairment on balance and visual processing functions in students with special educational needs"

Supplementary table 1. Comparison of demographic, visual and functional performance among subtypes of SEN [median (IQR) or percentage]

|  | CP† | ASD† | DS† | ID† | Others‡ | p-value |
| --- | --- | --- | --- | --- | --- | --- |
|  | (n = 18) | (n = 13) | (n = 12) | (n = 48) | (n = 13) |  |
| Demographic: |  |  |  |  |  |  |
| Chronological age (in years) | 16.0 (8.8) | 13.0 (7.5) | 17.5 (3.5) | 17.0 (4.8) | **12.0 (3.5)** | **< 0.01** |
| Sex  Female | 8 (44.4%) | 3 (23.1%) | 5 (41.7%) | 20 (41.7%) | 7 (53.8%) | 0.61 |
| Gestation age  Full-term (≥ 37 weeks)  Pre-term (< 37 weeks)  Unknown | 9 (50.0%)  9 (50.0%)  0 (0%) | 12 (92.3%)  0 (0%)  1 (7.7%) | 6 (50.0%)  2 (16.7%)  4 (33.3%) | 33 (68.8%)  4 (8.3%)  11 (22.9%) | 10 (76.9%)  2 (15.4%)  1 (7.7%) | **0.001** |
| Self-reported visual disabilities | **11 (61.1%)** | 2 (15.4%) | 3 (25.0%) | 13 (27.1%) | 2 (15.4%) | **0.02** |
| Vision measures: |  |  |  |  |  |  |
| Distance acuity of the better eye (LogMAR) | 0.83 (0.73) | 0.50 (0.73) | 0.48 (0.38) | 0.51 (0.66) | 0.51 (0.25) | 0.33 |
| Near acuity (LogMAR) | 0.40 (0.99) | 0.27 (0.64) | 0.45 (0.39) | 0.46 (0.73) | **0.20 (0.19)** | **< 0.01** |
| Contrast sensitivity (LogCS) | 1.64 (0.33) | 1.90 (0.46) | 1.60 (0.42) | 1.60 (0.80) | 1.60 (0.08) | 0.39 |
| Vision groups  Normal vision (NV)  Visual impairment (VI) | 4 (22.2%)  14 (77.8%) | 6 (46.2%)  7 (53.8%) | 6 (50.0%)  6 (50.0%) | 22 (45.8%)  26 (54.2%) | 4 (30.8%)  9 (69.2%) | 0.38 |
| Functional measures: |  |  |  |  |  |  |
| Visual orientation – time (s) | 1.58 (1.44) | 1.94 (1.38) | 1.90 (1.79) | 1.91 (1.43) | **1.18 (0.62)** | **0.04** |
| Facial expression (Cards) – time (s) | 20.77 (26.38) | 13.33 (16.21) | 11.75 (7.21) | 11.00 (17.75) | **2.33 (3.63)** | **0.001** |
| Facial expression (Examiner)– time (s) | 34.00 (48.19) | 17.00 (100.33) | 29.00 (37.25) | 17.00 (26.17) | **4.60 (6.35)** | **0.001** |
| Time-up-go (TUG) – time (s) | 15.36 (8.91) | 10.60 (3.80) | 17.69 (10.18) | 13.22 (7.39) | **9.03 (1.91)** | **0.001** |
| † CP: Cerebral palsy; ASD: Autism spectrum disorder; DS: Down syndrome; ID: Intellectual disability  ‡ Others special educational needs include deaf-mute, Rett syndrome, inborn errors of metabolism, etc. | | | | | | |
